# Donor Bone Marrow Derived Macrophage Engraftment into the Central Nervous System of Allogeneic Transplant Patients

**DOI:** 10.1101/2023.04.20.23288326

**Authors:** Anisha M. Loeb, Siobhan S. Pattwell, Soheil Meshinchi, Antonio Bedalov, Keith R. Loeb

## Abstract

Hematopoietic stem cell transplantation is a well known treatment of hematologic malignancies wherein nascent stem cells provide a regenerating marrow and immunotherapy against the tumor. The progeny of hematopoietic stem cells also populate a wide spectrum of tissues, including the brain, as bone marrow derived macrophages similar to microglial cells. We developed a sensitive and novel combined IHC and XY FISH assay to detect, quantify and characterize donor cells in the cerebral cortex of 19 female allogeneic stem cell transplant patients. We show that the number of male donor cells ranged from 0.14-3.0% of total cells or 1.2-25% of microglial cells. Using tyramide based fluorescent IHC we found at least 80% of the donor cells express the microglial marker Iba1 consistent with being bone marrow derived macrophages. The percentage of donor cells was related to pretransplant conditioning; donor cells from radiation based myeloablative cases averaged 8.1% of microglial cells, while those from non-myeloablative cases averaged only 1.3%. The number of donor cells in patients conditioned with Busulfan or Treosulfan based myeloablation were similar to TBI based conditioning; donor cells averaged 6.8% of microglial cells. Notably, patients who received multiple transplants and those with the longest post-transplant survival had the highest level of donor engraftment, with donor cells averaging 16.3% of microglial cells. Our work represents the largest study characterizing bone marrow-derived macrophages in post-transplant patients. The efficiency of engraftment observed in our study warrants future research on microglial replacement as a therapeutic option for disorders of the central nervous system.

**Key Points:**

1. We developed a sensitive Iba1 IHC and XY FISH co-stain assay to detect bone marrow derived donor cells in the brain of HSCT patients.
2. Donor cells in the CNS express Iba1 and average 8% of microglia after myeloablative HSCT; double transplants increase engraftment up to 25%.

## Introduction

Hematopoietic stem cell transplantation (HSCT), a form of cellular therapy used to treat hematologic malignancies and systemic genetic syndromes, provides both a nascent hematopoietic system and an immune system that includes donor-derived tissue resident macrophages. These cells engraft into a variety of tissues, including the brain, where they are called bone marrow derived macrophages (BMDM) and comprise a distinct population that shares many features of endogenous microglia.^1,2^

Microglial cells are unique among tissue-resident macrophages; they are exclusively derived from the yolk sac and are maintained by self-renewal.^3–6^ They function as immune sensors within the CNS and serve essential homeostatic functions in synaptic remodeling, antigen presentation, cytokine mediated immunity, neurogenesis and clearance of dead cells and debris.^3,6^ The functional capacity of BMDMs within the CNS and the extent of engraftment under normal conditions without injury or conditioning, are active areas of investigation. Studies in mice have shown that BMDM can functionally replace endogenous CNS microglia, treat genetic brain disease and deliver therapeutic proteins into the CNS.^7^ Studies with animal models have shown that microglial replacement therapy with BMDM is also an effective therapeutic strategy in a number of inherited metabolic diseases, microgliopathies, neuroinflammatory and neurodegenerative diseases including Alzheimer’s and Parkinson’s disease.^8–14^ In fact, there are multiple ongoing clinical trials to evaluate the therapeutic effect of microglial replacement *via* bone marrow transplantation for these disorders in humans^15–17^ and listed on www.ClinicalTrials.gov.

The ability of bone marrow-derived cells to stably engraft into the central nervous system (CNS) was best demonstrated in animal models transplanted with green fluorescent protein (GFP)-labeled hematopoietic stem cells.^18–20^ The engrafted GFP+ donor cells expressed the microglial marker Iba1 indicating they were BMDM.^18^ CNS engraftment was shown to require cranial irradiation as a conditioning agent; subsequent studies, however, demonstrated that Busulfan-based conditioning as well as immune or genetic ablation of endogenous microglial cells can also promote CNS engraftment.^21–24^ In fact, recent studies have reported near complete microglial replacement by using a combination of immune ablation and cranial irradiation in animal models.^23,25^

Donor-derived microglial replacement therapy (MRT) has been well studied in animal models, however, much less is known about BMDM engraftment in humans. Rare studies have reported the identification of a small number of donor-derived cells within the brain of a limited number of sex-mismatched stem-cell transplant patients.^26–28^ The dearth in human data regarding donor BMDM is attributed to the difficulties in performing XY FISH and IHC studies on poorly preserved postmortem tissue from stem cell transplant patients. In this study, we developed a modified XY fluorescence *in-situ* hybridization (FISH) assay and image analysis to enumerate and characterize donor-derived cells in the frontal cortex of 19 sex-mismatched hematopoietic stem cell transplant (HSCT) patients-the largest human study performed thus far. We evaluated the effect of conditioning and other variables including, time from transplant and show that donor-derived cells comprise as much as 25% of the microglial population.

Staining for the microglial marker Iba1 revealed that donor-derived cells are positive consistent with being BMDMs. Our observation of stable and robust engraftment of donor BMDM in the human brain provides the basis for future clinical studies using microglial replacement as a therapeutic modality.

## Materials and Methods

### Clinical Samples

Human cerebral cortex biopsies from female patients with a history of sex mismatched HSCT were obtained from the Fred Hutchinson autopsy tissue repository. Cases were selected based on post-transplant survival, conditioning regimens, and the number of transplants. Sex-matched HSCT cases were included as controls. All specimens used in this study were obtained after written consent from patients and approval by the FHCC Institutional Review Board (protocol #1837). The study was conducted in accordance with the Declaration of Helsinki.

### Fluorescence in-situ hybridization

Brain tissue sections were stained using modified FISH techniques optimized for heavily fixed postmortem tissue, with increased proteolytic digestion to unmask the DNA and overnight hybridization with Vysis CEP X (DXZ1) Spectrum Green Probe and Vysis CEP Y (DYZ1) Spectrum Orange Probe in Vysis IntelliFISH Hybridization Buffer (Abbott Molecular Diagnostics Des Plaines, IL) to visualize the X and Y chromosomes. (See Supplemental data for details). Following overnight hybridization, slides were washed and mounted in Antifade Mounting Medium with DAPI (Vectashield H-1200, Vector Labs; Burlingame, CA). FISH labeled sections were imaged with 40x/0.75 EC Plan-NEOFLUAR air objective on a Zeiss Axio Imager Z2 microscope as part of a TissueFAXS system (TissueGnostics; Vienna, Austria). Maximum projections of z-stacks (total size 7.2 μm, 0.8 μm interval) were used for image analysis.

### Donor cell quantification by XY FISH analysis

Scanned images were analyzed using TissueQuest (TissueGnostics, Vienna, Austria). DAPI stained nuclei were segmented by size to create nuclear masks. Background thresholds were defined to detect Y probe (red) and X probe (green) FISH signals within the nuclear mask.

A small sample region (5mm^2^ around 1,000 cells) was analyzed initially to ensure that automated detection using TissueQuest yielded accurate and comparable data as manual quantification. Following this validation, the number of donor and FISH positive cells were quantified in a larger area – 30-40mm^2^ encompassing 10,000-15,000 cells. Identified donor cells were also confirmed by blinded manual inspection. The percentage of donor cells was defined as the number of cells with a Y chromosome divided by the number of FISH positive cells (X or Y FISH signal). Donor cell quantification is presumably under-estimated since some donor cells were not identified because the tissue section did not include the Y chromosome (partial sampling of the nucleus). On average, approximately 66% of cells had a positive FISH signal (X or Y); 33% had a single X or Y signal and 33% had two signals (XX or XY).

### Combined Iba1 IHC and XY FISH staining

Selected cases were co-stained with Iba1 IHC and XY FISH to characterize donor cells. Sections were first incubated with anti-IBA1 antibody (Wako Chemicals, Japan), then with poly-HRP conjugated goat anti-rabbit IgG secondary antibody (Invitrogen; Carlsbad, CA), and visualized using Alexa Fluor 488 tyramide reagent (Invitrogen) per manufacturer instructions. The tyramide reagent forms a stable covalently ligated fluorochrome. The slides were then imaged by tissue fax. The same sections were then stained for XY FISH (as previously described) and reimaged by tissue fax. Since some of the IHC signal was lost following FISH, digital scans of the Iba1 IHC were overlayed onto FISH images utilizing DAPI stained nuclei for orientation (Adobe Photoshop).

### Immunohistochemistry (IHC)

Parallel sections of brain tissue were stained with anti-IBA1 antibody to detect and quantify microglia and bone marrow derived macrophages using standard conditions of citrate antigen retrieval and DAB detection (intelliPATH FLX, Biocare Medical). Iba1 positive microglia represented approximately 12% of the nucleated cells in the area used for XY FISH analysis.

## Results

The goal of our study was to detect and enumerate donor derived cells within the brain of stem cell transplant recipients. Brain tissue samples from female patients who had undergone a sex mismatched stem cell transplant were obtained from the Fred Hutchinson tissue repository. Cases were selected based on post-transplant survival, conditioning regiment, number of transplants, and absence of recent brain injury (Table 1). We focused on the frontal cortex since we observed reproducible staining and the highest number of donor cells in the cortical grey matter compared to sections from the pons and hippocampus.

**Table 1.**
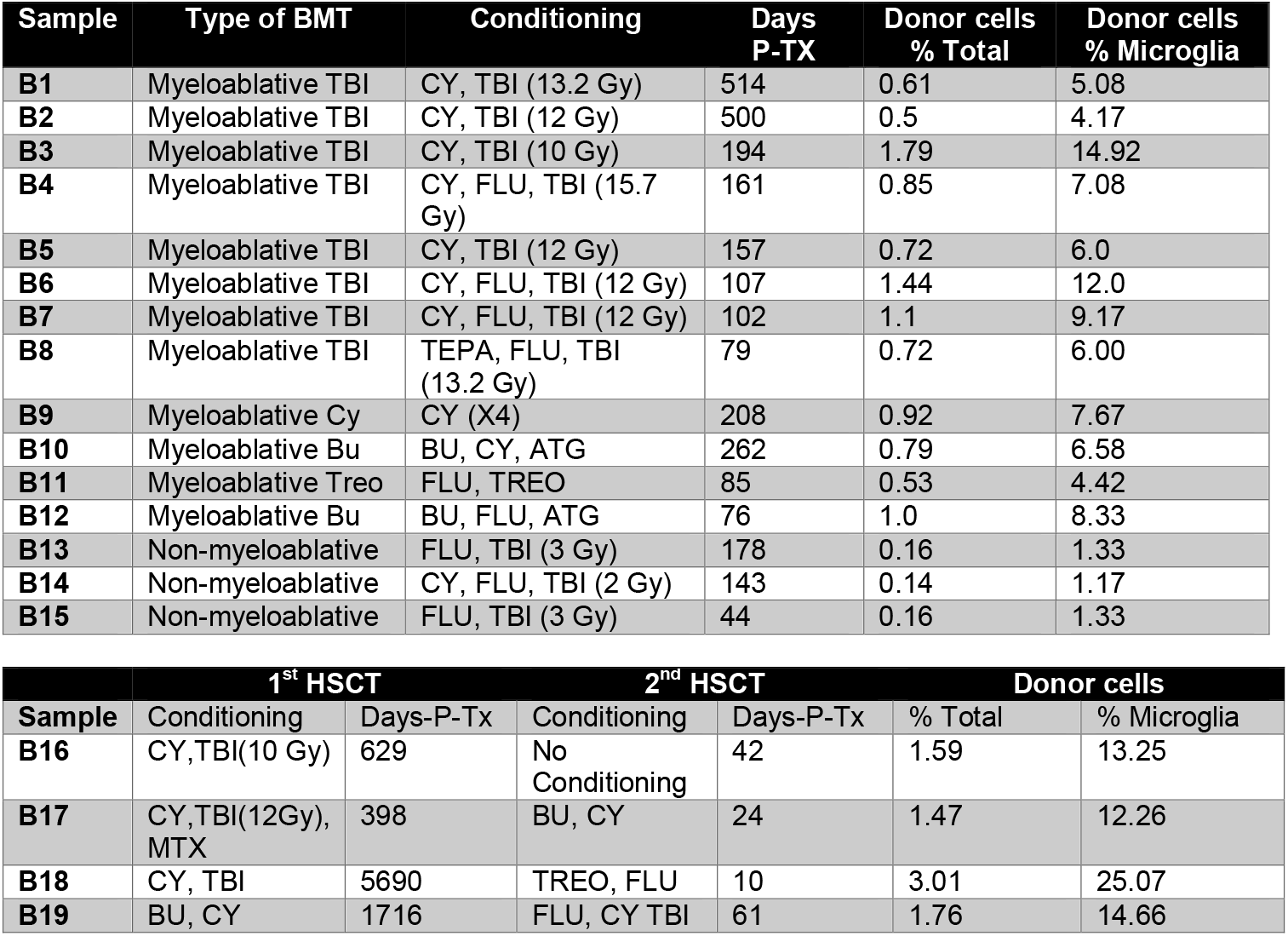
Patient cohort and conditioning regimens. Cerebral cortical biopsies were obtained from 19 female sex mismatched stem cell transplant patients. Cases were selected based on availability, conditioning regimen, post-transplant survival and number of stem cell transplants. B1-B8, patients conditioned with TBI based myeloablation (10-15.7 Gy); B9, patient treated with high dose (4X) Cytarabine myeloablation; B10-12 patients conditioned with Busulfan or Treosulfan based myeloablation; B13-15, patients conditioned with TBI based non-myeloablation; and B16-19, patients who received 2 separate stem cell transplants. Days-PTx, Post-transplant survival listed as days post-transplant. Donor cells are reported as percent total cellularity (donor cells % total) and percent of microglia cells (donor cell % microglia). TBI, total body irradiation; CY, Cytarabine; BU, Busulfan; TREO, Treosulfan; MTX, Methotrexate; FLU, Fludarabine; TEPA, Thiotepa; ATG, Antithymocyte globulin.

### Detection of donor cells by XY FISH

Fluorescence *in situ* hybridization (FISH) studies are difficult to perform on post-mortem brain tissue due to poor specimen preservation and background auto-fluorescence. To avoid these issues, prior studies have used chromogen-based detection, but with limited sensitivity. ^26,27^ We developed and optimized a more sensitive and robust XY FISH assay, which we utilized to detect male donor cells in cortex biopsies from female sex-mismatched stem cell transplant patients. The assay uses high concentrations of pepsin protease to unmask the DNA and commercial CEP Y spectrum orange probe and CEP X spectrum green Probe with IntelliFISH Hybridization Buffer to enhance the fluorescent signal. Using this assay, we detected male donor cells with XY staining within the cortical brain tissue-representative examples from 4 separate patients are presented in Figures 1A-D. X and Y chromosome detection was specific since no sex-mismatched cells were identified in tissue from female and male autologous transplant patients (Supplemental Figure 1). As is common with FISH-stained sections, only a single fluorescent signal (X or Y) was detected in many cells (50% of FISH positive cells), attributed to partial nuclear sampling in the thin tissue sections. Background autofluorescence was used to visualize tissue architecture including blood vessels, ventricles, and arachnoid membranes. We observed that most intraparenchymal donor cells existed as isolated cells enriched in perivascular regions (Figure 2A, B). Intravascular and peri-arachnoid donor cells were not included in the quantification but served as endogenous positive controls (Figure 2C, D). Additionally, although brain sections with infection, inflammation or adjacent hemorrhage had increased number of donor cells, they were excluded from the analysis.

**Figure 1.**
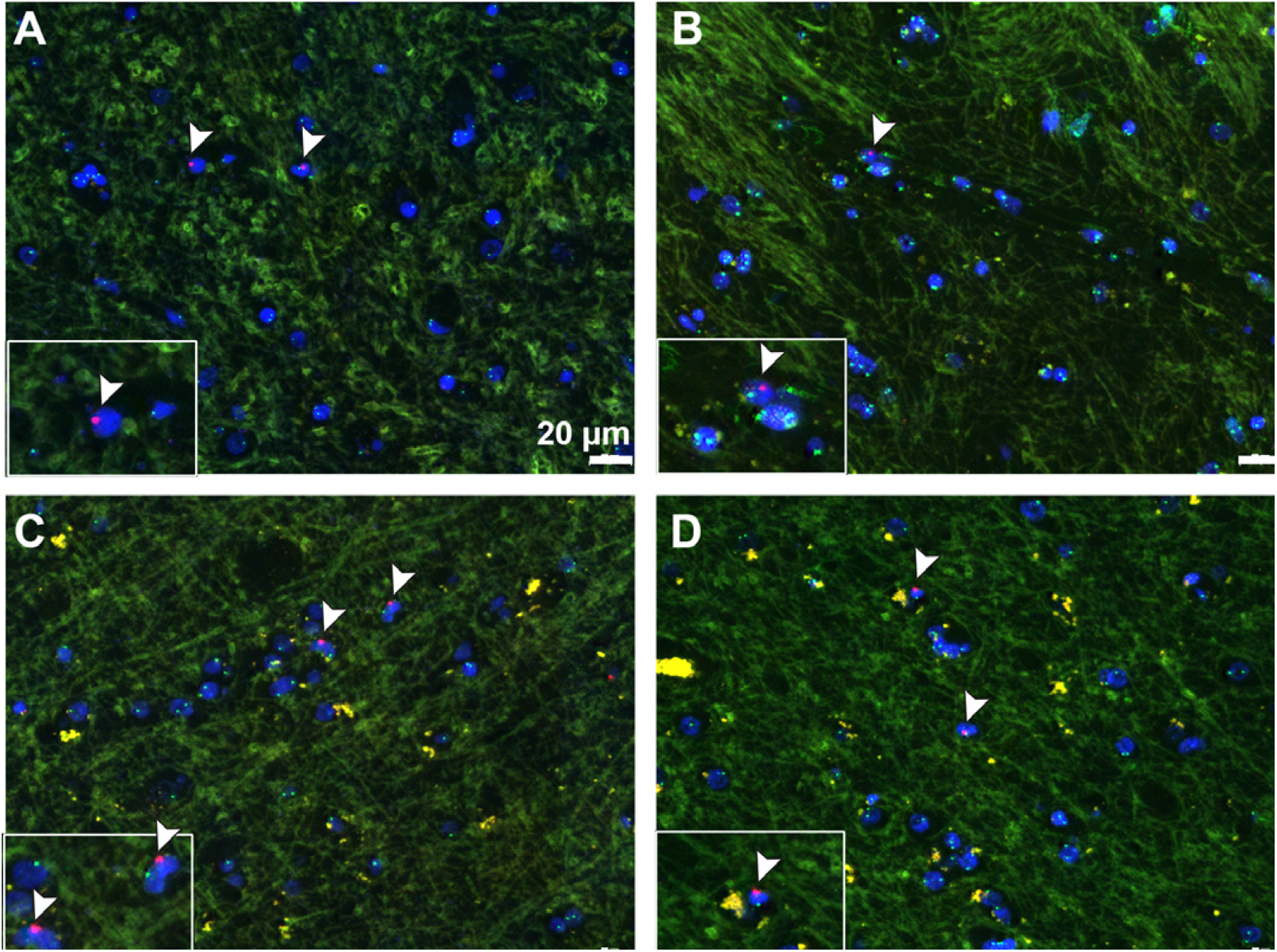
Detection of male donor cells by XY FISH. Representative XY FISH of frontal cortex biopsies from 4 separate female patients with history of sex mismatched stem cell transplant (A-D) Arrow-head, male donor cells. Light green, autofluorescence background highlighting tissue architecture. Red probe, Y chromosome; Green probe, X chromosome; Blue, DAPI nuclei. Original image at 400X magnification, Scale bar = 20μm.

**Figure 2.**
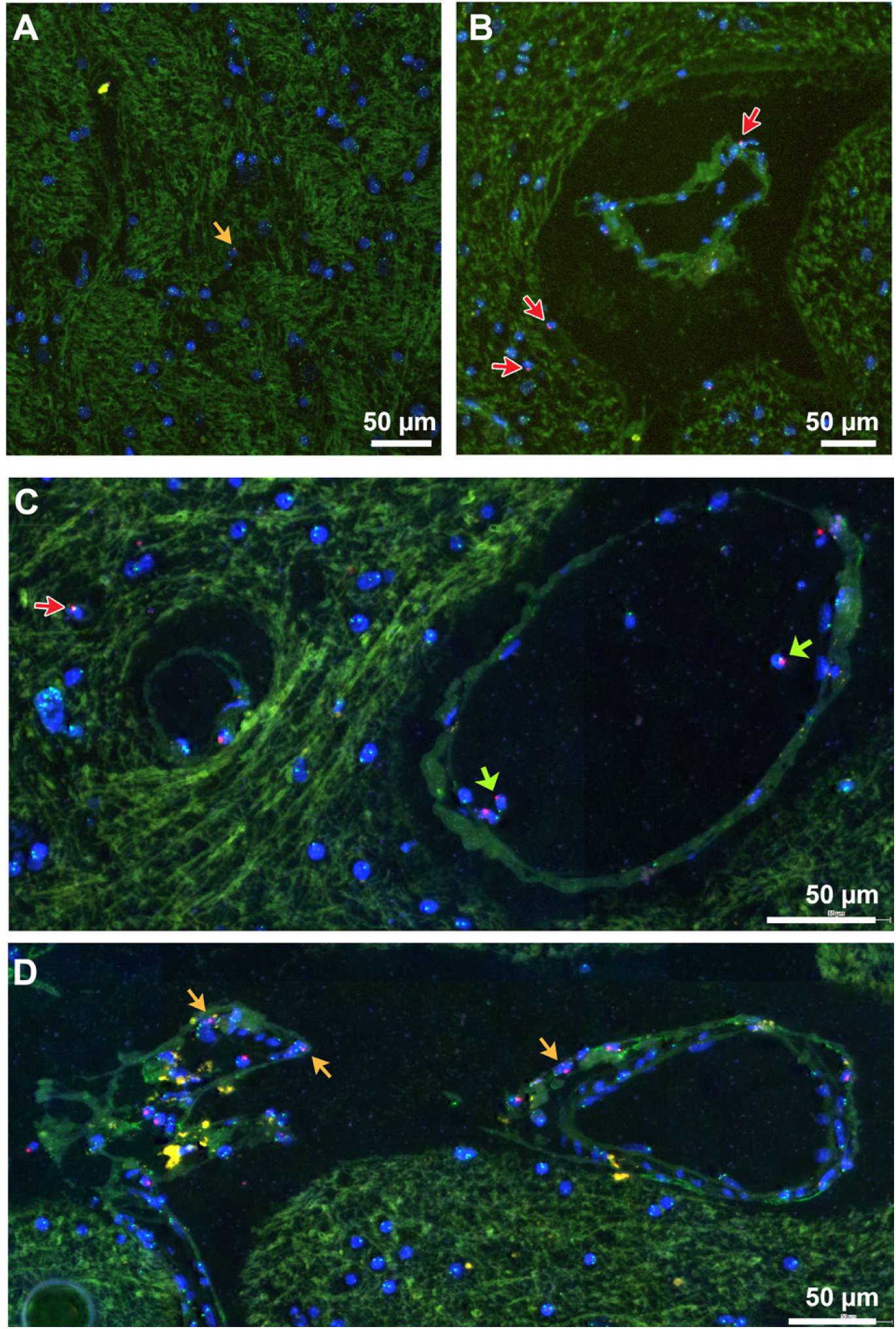
Distribution of male donor cells in cerebral cortex by XY FISH. Representative cortical biopsy sections from female patients with history of sex mismatched stem cell transplant illustrating distribution of male donor cells. (A) Most donor cells exist as individual cells within the cortical gray mater (orange arrow). (B&C). Male donor cells enriched in perivascular location (red arrows). Background autofluorescence identifies small blood vessels and intravascular donor cells (green arrow). (D) Donor cells are enriched along cortical surface adjacent to arachnoid membrane (orange arrows). Red probe,Y chromosome; Green Probe, X chromosome; Blue, DAPI stained nuclei. Original image at 400X, scale bar = 50μm.

### Characterization of donor cells

Most CNS engrafted donor-derived cells in animal models are thought to be BMDM based on the expression of Iba1.^18^ To confirm that the donor cells we detected in our patient samples are in fact BMDM, we established a novel combined Iba1-IHC and XY-FISH assay. Since protease treatment required for FISH staining eliminates the Iba1 epitope, and stable 3,3′-Diaminobenzidine (DAB) IHC blocks the fluorescent FISH signal, we developed a tyramide-based fluorescent IHC assay in which a covalent ligated fluorochrome is formed and preserved during subsequent FISH staining. Tissue sections were first stained for Iba1, scanned, and then stained for XY FISH followed by rescanning on the TissueFAXS. Since some of the IHC fluorescent signal is still lost following FISH, scans of Iba1-IHC stained sections were overlayed onto XY-FISH images using DAPI stained nuclei for orientation (Figure 3). The overlapped images showed that most donor cells (>80%) express Iba1, consistent with having BMDM identity (Figure 3). Additionally, similar to endogenous microglia, Iba1 positive donor cells were present in both ramified and ameboid forms (Figure 3, arrows) with ameboid cells (upper panel) having higher levels of Iba1 than the ramified forms (lower panel).

**Figure 3.**
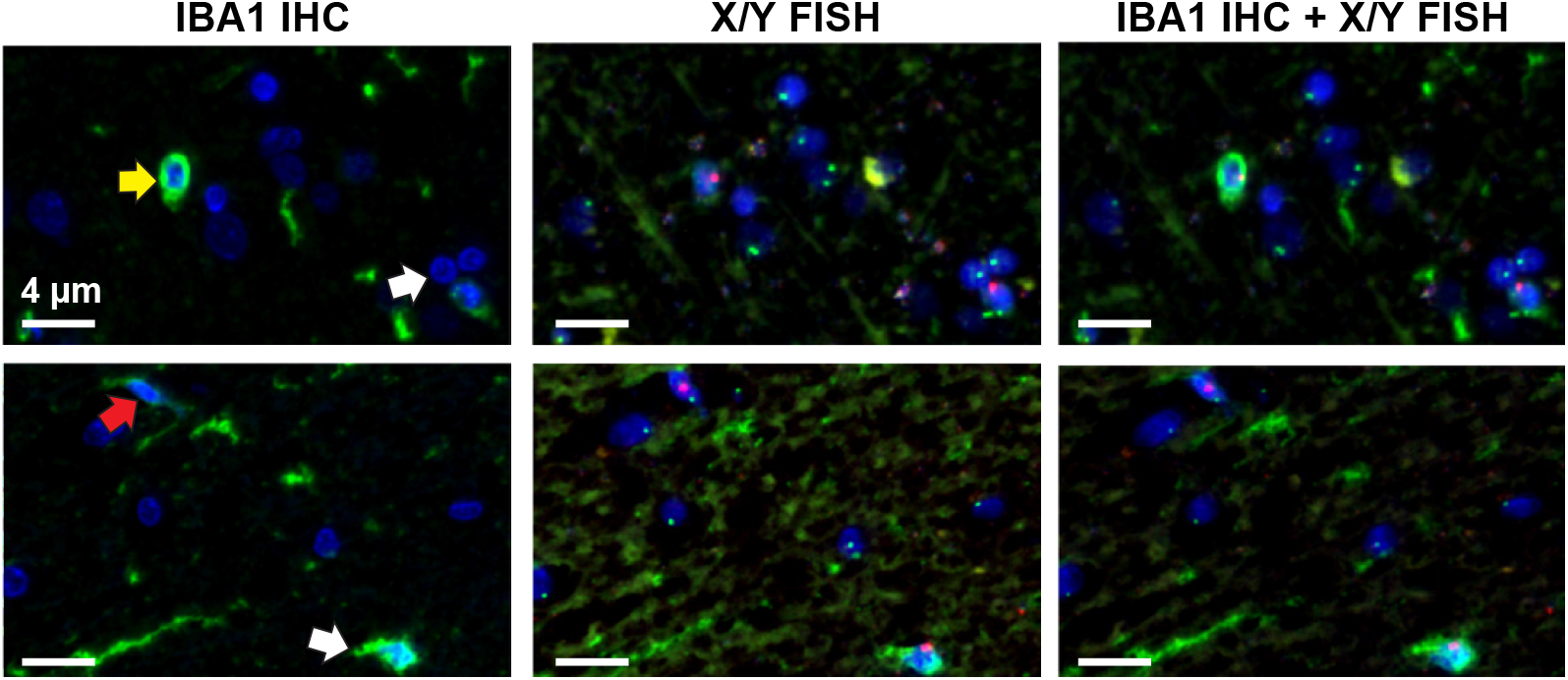
Tyramide IHC and XY FISH show male donor cells express Iba1. Representative images from tyramide Iba1 IHC and XY FISH from two separate samples (top and bottom rows). (Column 1) tyramide based Iba1 IHC (Green); (Column 2) same sections following XY FISH; (Column 3) digital overlap of IHC and FISH images. Yellow arrow (top) and red arrow (bottom), donor cells. Donor cells are present in both amoeboid (yellow arrow) and ramified (red arrow) forms with lower expression of Iba1 in the ramified cells. Green, Iba1 IHC; Red probe, Y chromosome; Green probe, X chromosome; Blue, DAPI stained nuclei. Images taken at 400X original magnification, scale bar = 4μm.

### Quantification of donor cells

To quantify donor derived cells, we digitally scanned the slides using a tissue fax and then set up conditions to detect green X and red Y probes within DAPI stained nuclei. The automated detection system efficiently identified donor cells based on the presence of the Y chromosome (red probe). However, variable autofluorescence and background staining hindered automated detection of the X chromosome (green probe). We first calibrated image analysis parameters in a small area that was then compared to manual evaluation. The established parameters were then applied to a larger region encompassing approximately 10,000-15,000 DAPI positive cells. Identified donor cells were confirmed by blinded manual inspection. We defined the percentage of male donor cells as the number of cells staining with the Y probe compared to total FISH positive cells. Cells without detectable FISH signal were excluded from our analysis.

### Effect of Conditioning Regimens on donor cell engraftment

Preclinical studies have shown that host microglial depletion or pre-transplant conditioning that disrupts the blood-brain barrier, facilitate donor cell engraftment into the brain.^29,30^ In agreement, we found that pretransplant conditioning had a significant effect on donor cell engraftment into the CNS. Donor cells were quantified as described in Material and Methods. Male donor cells comprised an average of 8.1% (4.2-14.9%) of the microglial population following myeloablative conditioning (total body irradiation (TBI) >1000 cGy) while in non-myeloablative cases (TBI<300cGy), they averaged only 1.3% (Figure 4A, Table 1). The percentage of donor cells relative to the microglial population was calculated based on microglial cells representing 12% cellularity within the cortical gray matter as determined from independent Iba1 IHC studies (Figure 5) in agreement with other studies.^31^ We also included cases with Busulfan or Treosulfan-based myeloablative conditioning and found that both regimens supported BMDM engraftment, with donor cells averaging 6.8% (4.4-8.3%) of microglia (Figure 4A, Table 1). Since donor cell engraftment maybe limited by the low numbers of circulating donor precursor cells (monocytes) in the early post-transplant period, repeat stem cell transplants could increase the number of donor cells in the CNS.^32^ Consistently, we observed that patients with 2 or more transplants showed higher levels of donor cell engraftment than those with a single transplant, averaging 16.3% (12.2-25.1%) of microglial cells (Figure 4A; Table 1 B16-B19) Preclinical studies have reported that engrafted BMDM are long-lived and capable of self-replication with clonal donor cell numbers increasing over time following transplant.^25,33^ Our data agree with these reports. We observed that donor cell number increases and correlates with post-transplant time (Figure 4B). The patient who survived the longest (>15 years post-transplant) had the highest number of male donor cells (25% of the microglial population; (Figure 4B; Table 1). However, since this patient also received multiple transplants, the higher number of donor cells could be due to longer post-transplant survival, repeat stem cell transplants, or a combination of the two.

**Figure 4.**
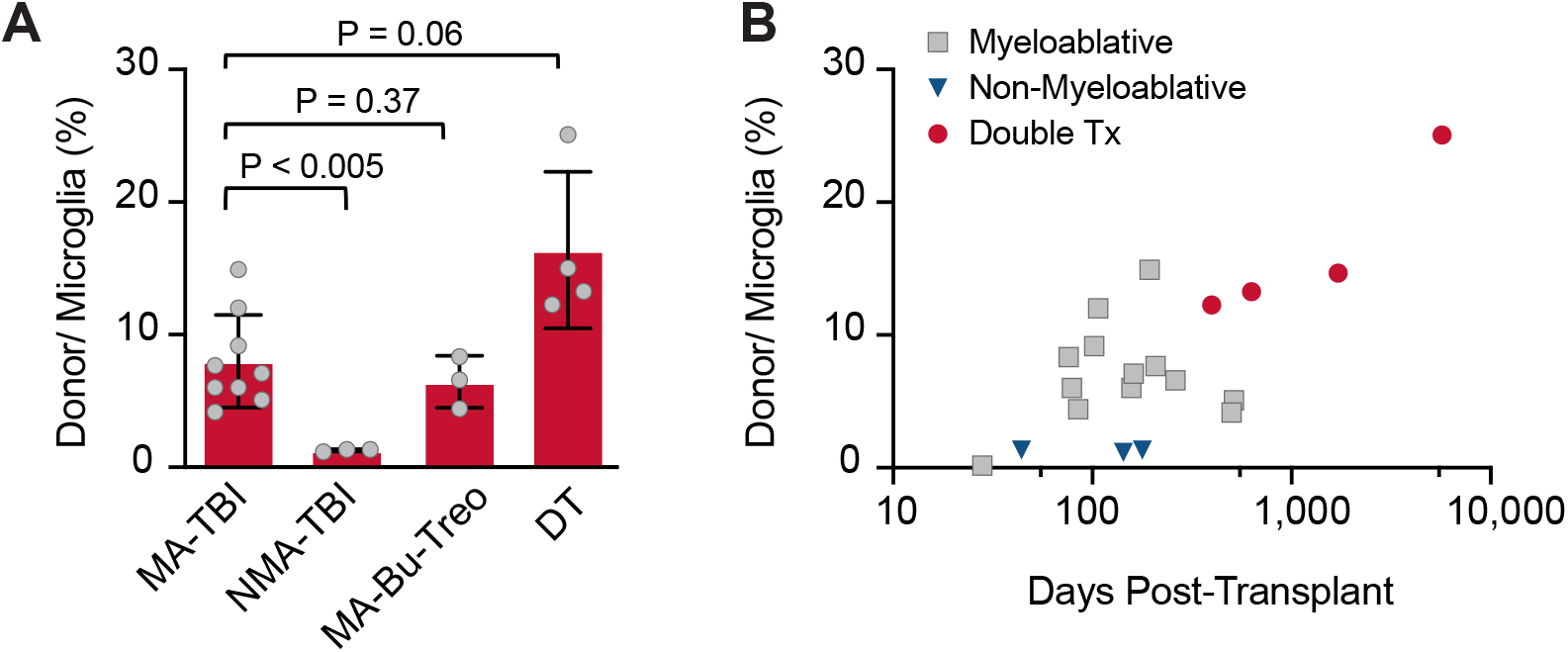
Effect of pretransplant conditioning on donor cell engraftment in cerebral cortex. (A) Percentage of male donor cells identified in cerebral cortex biopsies from patients conditioned with total body irradiation (TBI) based myeloablation (MA-TBI; total body irradiation >1000 cGy), TBI based non-myeloablation (NMA-TBI; Non-myeloablative conditioning, TBI<300cGy), Busulfan or Treosulfan based myeloablation (MA-Bu-Treo) or with two separate stem cell transplants (DT; double transplants). Data are expressed as donor cell percentage relative to total microglial cells. Microglia cells represent approximately 12% of cellularity in cerebral cortex based on parallel Iba1 IHC studies (supplemental Figure 2). (B) Effect of post-transplant survival on donor cell engraftment in cerebral cortex. Data are expressed as percentage of donor cells relative to total microglial cells. Gray square, myeloablative transplant; blue inverted triangle, non-myeloablative transplant, red circle, double transplants.

## Discussion

Pre-clinical studies have demonstrated the potential of microglial replacement therapy as a form of cellular therapy to treat inherited brain disease and deliver therapeutic proteins into the CNS.^7,16^ Although MRT is well studied in murine models there are very only very few studies documenting donor derived cells in the brain of stem cell transplant patients. Here, we present the largest and most comprehensive study, using a cohort of 19 female patients with a history of sex mismatched stem cell transplant, to identify and characterize donor derived cells in the cerebral cortex. Using a sensitive XY FISH assay optimized for post-mortem tissue, we show that donor cells represent approximately 1% of the total and 8.4% of the microglia population, respectively. Co-staining with modified IHC and FISH showed that most donor cells express the microglial marker Iba1 indicating they are bone marrow derived macrophages akin to endogenous microglia. The percentage of donor cells is remarkably conserved among patients with similar pre-transplant conditioning. Pre-transplant conditioning strongly influences donor cell engraftment, with patients receiving myeloablative transplants having much higher numbers of donor cells than those with non-myeloablative transplants. In agreement with animal studies, we show that Busulfan or Treosulfan-based conditioning is sufficient to support bone marrow macrophage engraftment at similar levels as those with other myeloablative regimens. The engrafted cells are stable over the post-transplant period and trend to increase with post-transplant survival. Finally, patients who received two or more stem cell transplants had the highest number of donor derived cells, which may either be related to repeat transplants or the long post-transplant survival period.

Microglial replacement therapy has great promise for the treatment of a number of CNS disorders.^25^ Preclinical studies have shown that wild type or genetically modified cells can be used to treat a number of microgliopathies including of metabolic disorders such as adrenoleukodystrophy^34^, lysosomal storage diseases including Hurler syndrome^16,17,35^, as well as neuroinflammatory and neurodegenerative disease including ALS^36^, Alzheimer’s^10,11,37^ and Parkinson’s disease.^9^ In these disorders, the donor bone marrow derived macrophages partially replace the defective microglia, can supplement normal microglial functions as well as provide needed wild type proteins. Microglia replacement therapy can also be used as a therapeutic strategy to replace defective microglia in CSF1R-related leukoencephalopathy.^16,38^ Of interest, we showed that Acute Myeloid Leukemia patients that express a polymorphism of CD33 are resistant to gemtuzamab (an antibody drug conjugate directed again CD33)^39^ and genetic studies have shown this same polymorphism is protective against Alzheimer’s disease.^40,41^ This has led us and others to propose that autologous MRT with modified stem cells expressing the CD33 isoform may delay or prevent Alzheimer’s disease.^37,42^

There are several active clinical studies involving microglia replacement listed on www.ClinicalTrials.gov. However, in many instances significant neurologic improvement has been variable and limited.^17,43^ The lack of therapeutic efficacy in clinical trials involving MRT may be related to suboptimal donor cell engraftment and/or treatment of patients with advanced disease. Further, many of these CNS disorders are complex and require early therapeutic intervention prior to irreversible tissue and neuronal damage. In addition, cognitive and synaptic defects in mice with long-term colonization of marrow derived macrophages in the CNS has also been reported.^44^ Despite these limitations, the future of MRT looks promising with recent advances in minimizing transplant toxicity, enhancing the efficiency of cellular engraftment, and using genetically modified donor stem cells. For example, a combination of pre-transplant microglial depletion *via* CSF1R inhibition and conditioning has yielded >92% donor cell engraftment in murine models of MRT.^23^ Another study reported that post-transplant CSF1R inhibition resulted in near complete BMDM engraftment with therapeutic efficacy in a mouse model of Prosaposin deficiency.^25^ In contrast we report engraftment rates as high as 25% with human brain samples. It will be interesting to see whether the above approaches can be employed to increase the efficiency of MRT even further to provide long-term therapy for intractable neurologic disorders in humans.

## Data Availability

All data produced in the present study are available upon reasonable request to the authors

## Acknowledgements

This study was supported by grants from Core Center of Excellence in Hematology Grant (SSP +KRL), Seattle Translational Tumor Research (STTR) Data Generation and Bioinformatics Grant (KRL) and Rett Syndrome Research Trust (AB). The authors would like to thank Lena Schroeder and Dave McDonald, Cellular Imaging Shared Resource RRID:SCR_022609 of the Fred Hutch/University of Washington/Seattle Children’s Cancer Consortium (P30 CA015704). Authors would like to thank Diana Lim for help with images and figures; Andy Larson, Brandon Seaton, Kristin Shrimp and histology lab, Fred Hutch Cancer Center; and Lena Glaskova, Fred Hutch Cytogenetics lab.

## Authorship Contributions

Contributions: KRL designed the experiments; AML and KRL wrote the paper; AML, SSP and KRL performed the experiments and analyzed the data; SM, SSP and AB provided general scientific guidance and designed the experiments. All authors reviewed the manuscript prior to submission.

