## supplemental figures for "Donor Bone Marrow Derived Macrophage Engraftment into the Central Nervous System of Allogeneic Transplant Patients"

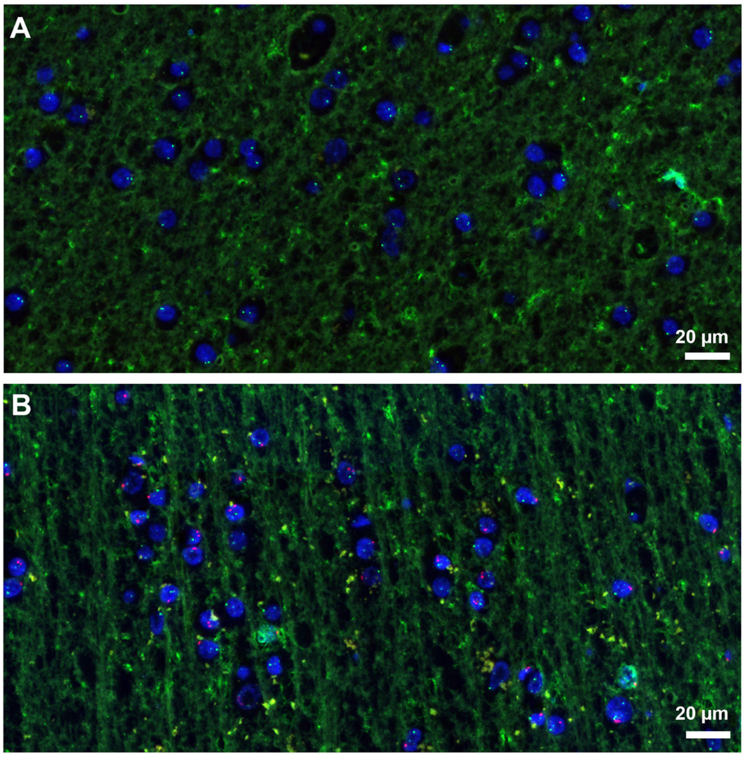


Supplement Figure 1
K. Loeb, et al.

**Supplemental Figure 1. Control XY FISH on autologous stem cell transplant samples.**
XY FISH of frontal cortex biopsies from female (A) and male (B) autologous stem cell transplant patients/recipients. No definitive sex mismatched cells were detected. Some cells showed staining for only a single probe, hindering detection of possible female cells in the male autologous transplant sample (B). Original magnification 200x; scale bar = 20 mm.
Green probe, X chromosome; Red probe, Y chromosome; Blue DAPI stained nuclei.

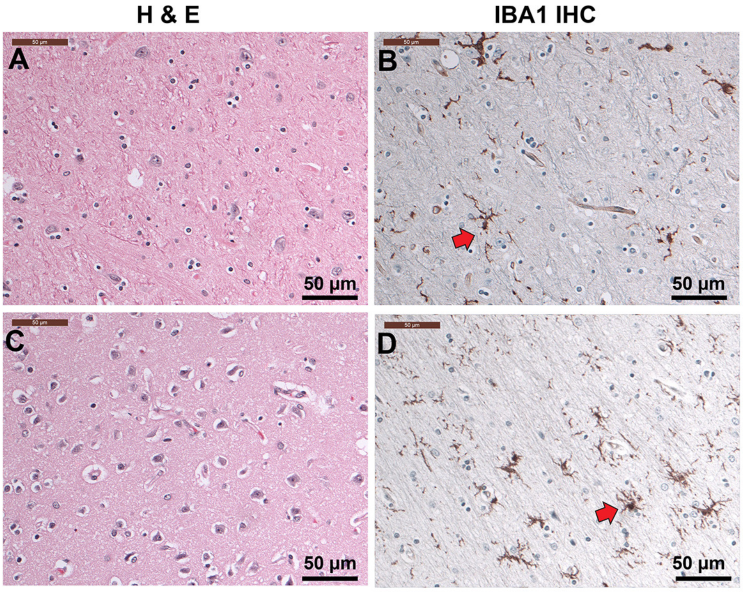


Supplement Figure 2
K. Loeb, et al.

**Supplemental Figure 2. H&E and Iba1 IHC stained frontal cortex sections.** Representative H&E-stained sections of cortical grey matter from post-transplant patients without morphologic abnormalities (A+C). Iba1 IHC stained sections of cortical grey matter (B+D) show that microglia cells represent approximately 12% of nucleated cells. Most of the microglial cells are present in ramified or resting forms (B+D) with long cellular processes (arrow). Original image 200x; scale bar = 50 mm.
